# Predicting elimination of evolving virus variants

**DOI:** 10.1101/2021.06.24.21259501

**Authors:** Elliott Hughes, Rachelle Binny, Shaun Hendy, Alex James

## Abstract

As the SARS-CoV-2 virus spreads around the world new variants are appearing regularly. Although some countries have achieved very swift and successful vaccination campaigns, on a global scale the vast majority of the population is unvaccinated and new variants are proving more resistant to the current set of vaccines. We present a simple model of disease spread which includes the evolution of new variants and varying vaccine effectiveness to these new strains. We show that rapid vaccine updates to target new strains are more effective than slow updates and containing spread through non-pharmaceutical interventions is vital whilst these vaccines are delivered. Finally when measuring the key model inputs, e.g. the rate at which new mutations and variants of concern emerge, is difficult we show how an observable model output, the number of new variants which have been seen, is strongly correlated with the probability the virus is eliminated.

## Introduction

The recent emergence of multiple SARS-CoV-2 variants poses a risk to the global efforts to control the COVID-19 pandemic. In the short term the increased transmissibility of some variants raises the risk of uncontrolled outbreaks (Davies et al., 2021; Tegally et al., 2020). In the long term, the increasing genetic diversity of the virus may result in reduced vaccine efficacy due to immune escape, i.e. variants may mutate so that antibodies which target other variants cannot successfully bond to viral particles from the new variant (Atlani-Duault, Lina, Chauvin, Delfraissy, & Malvy, 2021; Callaway, 2021). Many models for COVID-19 spread have assumed that only one variant will be active at any given time (James, Plank, Binny, Hannah, et al., 2020; James, Plank, Binny, Lustig, et al., 2020), a simplifying assumption which is no longer accurate. Here we construct a simple stochastic branching process model for multi-strain epidemics of COVID-19. By simulating outbreaks under different assumptions, we suggest some possible dynamics for genetic diversity and propose observable measures to assess the effect of these multiple strains on long term elimination.

Genomic data of the Sars-CoV-2 virus has been widely shared worldwide. Although RNA viruses in general tend to mutate quickly, coronaviruses in particular mutate much slower, likely due to a proof-reading enzyme that corrects copying mistakes which are potentially fatal to the virus. Although 12,000 mutations were spotted in Sars-CoV-2 within a year of its emergence almost all of these appeared to have almost no consequence on the virus’s transmission rate (Callaway, 2020).

The mutation rate of a virus can be thought of on many levels. At the fastest level there are small genetic mutations which happen at almost every infection and can be used to track individual infection routes by public health departments. Conversely, there are the much-publicised Variants of Concern (VoCs), highly significant mutations seen across many individuals (Centers for Disease Control and Prevention (CDC), 2021; World Health Organization, 2021). For a new strain to be classified as a Variant of Concern there must be evidence of impact on diagnosis, treatments or vaccines, increased transmissibility or increased severe disease. Our model includes only impact on vaccines and increased transmissibility, and like reality not all new variants that appear in the model will have these characteristics, most variants are simply small changes of very little consequence. As of May 2020, the CDC had recognised five Variants of Concern (VoC) for Covid-19 which were all first detected in regions that had high infection rates (B.1.1.7 -UK, P.1 – Brazil, B.1.351 – South Africa, B.1.427 and B.1.429 – California, USA). Conversely there are many regions that have had very high infection rates and no VoCs have been recognised (e.g. France, Italy, Spain, Russia). This could be in part due to low rates of genetic sequencing but it is likely that no VoCs have arisen in many of these areas despite the high transmission rates.

Although the number of VoCs is small, the number of mutations seen in widespread genetic testing is much larger. Within a year of the virus emerging there were many distinct viral clades in global circulation (Hamed, Elkhatib, Khairalla, & Noreddin, 2021). However, it was after 13 months in Jan 2021 that the first VoC was identified, B.1.1.7, which rapidly became the main variant in circulation in Europe (Planas et al., 2021). The current literature on the genetic diversity of SARS-CoV-2 focuses on reconstructing past genetic trees rather than forecasting the long-term number of active antigenically distinct variants e.g. Nie et al. (2020). While these analyses provide useful information, as vaccines have begun to be distributed it has become crucial to determine the risk of an antigenically distinct variant emerging.

Multi-strain influenza outbreaks have been modelled using both branching processes and compartmental models (Kucharski, Andreasen, & Gog, 2016). Branching processes are inherently more computationally intensive than compartmental approaches and as a result they are more often used to find the probability of a new variant becoming established than to predict long-term genetic diversity e.g. Bedford, Rambaut, and Pascual (2012). Compartmental models can be broadly separated into ‘history-based’ and ‘status-based’ approaches. History-based models attempt to capture the infection history of each individual, while status-based models only record the current immune status of individuals. While history-based models contain more detail, a naïve history-based model for n strains will have at least 2*n* variables (Kucharski et al., 2016) and may be computationally intensive as a result. Various attempts have been made to reduce the number of variables necessary for a history-based model, but these either require certain assumptions to be met and/or have increased analytical complexity. Status-based models are simpler and have been used to model the dynamics of other RNA viruses, e.g. Koelle, Kamradt, and Pascual (2009) or Koelle, Khatri, Kamradt, and Kepler (2010), but often lack realistic details. For a summary of the advantages and disadvantages of various approaches, see Kucharski et al. (2016).

Table S1 gives details of the administration and uptake of vaccines in different countries, and the incidence of each VoC (as a proportion of all sequenced infected cases). As of early April all countries with a significant number of infections had significant numbers of infections by at least one VoC. Between March and April 2021 there was a clear rise in the number of infections caused by new variants of SARS-CoV-2, in particular the B.1.1.7, P.1 and B.1.617.1 variants. Vaccination rates as of April 2021 varied between <0.1% of the population daily, e.g. New Zealand, and over 0.8% of the population daily, e.g. UK and USA. The proportion of the population who are likely to be vaccinated ranges from less than 50% (USA) to over 75% (NZ, Brazil).

Our model tracks virus mutations at a rate somewhere between the two extremes, we assume there is a small probability the virus can mutate each time a new person is infected. Although in reality mutations happen within a single person our model assumes each individual has single variant which does not change over time and the resulting secondary case is infected by the new mutation. The mutation rate parameter is intended to track significant mutations where the transmission rate of the virus changes significantly (either increasing or decreasing) rather than each individual genetic mutation. A new variant will have a different transmission rate to the parent variant it evolved from but this rate may be smaller and hence the variant quickly vanishes.

The model includes selection pressure as new variants are more likely to gain traction and spread to many individuals if they have a higher transmission rate than other variants, and are able to evade any immunity the population has already built up from infection by or vaccination against other variants.

We explore the model and give results on how the probability of elimination changes as the speed at which vaccines are rolled out is increased for different cross-immunity and mutation rate scenarios. Finally, we discuss how to estimate the probability of elimination in a system where the key parameters are not known.

## Model

The model consists of four separate parts: disease transmission, virus evolution, immunity and vaccination. The disease transmission model is based on a simple branching process. As each infection occurs there is a small chance the virus mutates into a new variant. The virus evolution model builds a simple tree of these variants tracking how closely related they are. The immunity model predicts immunity to infection depending on how closely related the virus variants are. Finally we consider a number of immunisation strategies and assess their effect on the epidemic spread.

### Disease transmission model

We consider a population of *N*_*p*_ individuals. Each realisation starts with *N*_0_ infected individuals who expose, on average, *R*_1_ individuals each over *D*_*Inf*_ days. *N*_0_ is chosen to be sufficiently high such that in a fully-susceptible population almost all realisations will result in an epidemic wave rather than elimination by chance. New exposures to an infected individual occur uniformly over that individual’s infectious period of *D*_*Inf*_ days. The actual number of exposures caused by an infectious individual is Poisson distributed with expected value *R*. The population is well-mixed, i.e. all individuals are equally likely to come into contact and infect each other. An exposed individual will become infected provided they are not currently infected and are not immune to that variant of the virus. There is no latent period, once an individual is infected they start infecting others immediately. Individuals can acquire immunity (after infection or vaccination) so the number of susceptible individuals in the population varies over time, however the model assumes there are no fatalities so the population size *N*_*p*_ remains constant over time. Once an individual is immune to a particular strain they remain immune forever, i.e. there is no waning immunity.

### Virus evolution model

When an individual is infected they are usually infected with the same virus variant as the index case from whom they were infected. However, there is a small probability, *p*_*mutate*_, that they will be infected with a new variant of the virus. Each new variant has its own transmission rate, *R*_*j*_, which is the expected number of new cases caused by a single individual infected with variant *j* in a fully susceptible population, i.e. no individual has immunity to variant *j*. The transmission rate of a new variant, *R*_*j*_, depends on the transmission rate of the parent variant, *R*_*p*_

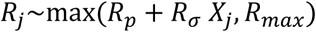

where *X*_*j*_ is a normally distributed random variable, *X*_*j*_∼*N*(0,1). We cap *R*_*j*_ at a maximum rate *R*_*max*_. This assumes that transmission rates will be self-limiting as social distancing measures imposed either by individuals or at a population level will stop very high transmission rates e.g. when transmission rates are high, non-pharmaceutical interventions are likely to be implemented to curb spread and public perception of risk increases.

Each new variant is a new node of the virus tree model and edges connect new variants with their parent variant. The model produces a non-binary, rooted tree which allows multiple edges from a single node as a variant begets many different mutations. The distance *d*_*ij*_ between nodes representing variants *i* and *j* tells us how closely genetically related the two variants are, e.g. if two variants share the same parent variant they are distance 2 apart on the virus tree.

### Immunity model

Once an individual has been infected with variant *j* of the virus they naturally acquire full immunity to that variant. However, they may still become infected by a different stain of the virus. The probability that an individual who has been infected with variant *j* is immune to variant *i* is *I*_*ji*_. *I*_*ji*_ is a decreasing function of, *d*_*ji*_, the distance between nodes *j* and *i* in the virus tree. If two variants are very closely related there is a high probability that infection by one variant will confer immunity to the other. If variants are more distantly related the probability is much lower. Parameters are chosen so that if two variants are more than distance 6 from each other on the tree then there is a zero probability of immunity.

Although the immunity matrix, *I*, gives a probability of being immune to a particular variant, a particular individual’s immunity to a given variant is a binary measure, i.e. if *I*_*ji*_ = 0.5 then 50% of individuals who have been infected with variant *j* are fully immune to variant *i* and 50% are fully susceptible. Immunity across variants is independent, e.g. catching variant A may make one person immune to variant B but not variant C, whereas it may confer immunity to both variants B and C in another individual.

### Vaccinations

If a vaccine is developed to give immunity to a particular variant then, like previous infection with that variant, it is presumed to be 100% effective against that variant after a single shot. It also confers partial immunity against other variants following the same rules as the natural immunity model. Vaccination begins on day *D*_1_ = 350 and fraction *f*_*v*_ individuals are vaccinated each day. When a vaccine is updated the old vaccine is discontinued immediately and all subsequent vaccinations use the vaccine developed against the new variant. Individuals are vaccinated in a priority order, those who have been infected are equally likely to be vaccinated as those that have not. Initially we use a Continuous vaccination strategy i.e. when the last person on the priority list has been vaccinated the list resets and returns to the first individual till either the entire population has received that vaccine or the vaccine is next updated.

The model is run in time steps of one day. Results are averages taken from 1000 simulations at each set of parameter values. All fixed parameter values are given in Table 1.

**Table 1:** Fixed parameter values.

| Description | Parameter |
| --- | --- |
| Population size | $N_p = 10,000$ |
| Initial number of infected individuals | $N_0 = 10$ |
| Initial variant transmission | $R_1 = 1.2$ |
| Maximum transmission<br>(high and low transmission) | High: $R_{max} = 1.8$<br>Low: $R_{max} = 1.4$ |
| Transmission variation of new variants | $R_\sigma = 0.1$ |
| Number of infectious days | $D_{Inf} = 14$ |
| Immunity rate at distance $d = 1, \dots, 6$<br>(high and low cross immunity) | High: $I(d) = 1, 0.9, 0.5, 0.3, 0.1, 0.05$<br>Low: $I(d) = 0.5, 0.45, 0.25, 0.15, 0.05, 0.025$ |
| Virus mutation rate | $p_{mutate} = 0.01$ |
| First day of vaccinations | $D_1 = 350$ |

## Results -No vaccines available

We start by considering results from the model assuming no vaccine is available. The raw model output is a time series of the number of individuals infected with each virus variant and the tree of the evolving virus. Each model realisation shows one of two distinct types of behaviour: either the disease is self-limiting, the population reaches herd immunity, all variants die out and the disease is eliminated; or the disease undergoes variant runaway where there are multiple variants in circulation, and the virus is never eliminated as new variants appear faster than herd immunity to each variant is achieved. This second situation is similar to the current global circulation of influenza viruses. Due to the stochasticity in the model it is possible to see both types of behaviour at a single set of parameter values.

Figure 1 shows example output for each of these behaviours. In both of these examples there is no vaccination and herd immunity can only be reached through infection. In the first realisation (Figure 1 A, C, E) after approximately 600 days the virus has been eliminated. There were 2 distinct waves of infection, peaking at days 150 and 440. Both peaks were dominated by a single virus variant though other variants were active. The number of active variants, i.e. variants with at least one infectious individual, peaks shortly after the peak in the number of infectious cases (Fig 1A). The virus tree shows that the original variant produced 13 new variants. It was the last of these that appeared around day 250 that dominated the second wave and produced 12 new variants. The size of the node shows the transmission rate of each variant.

**Figure 1:**
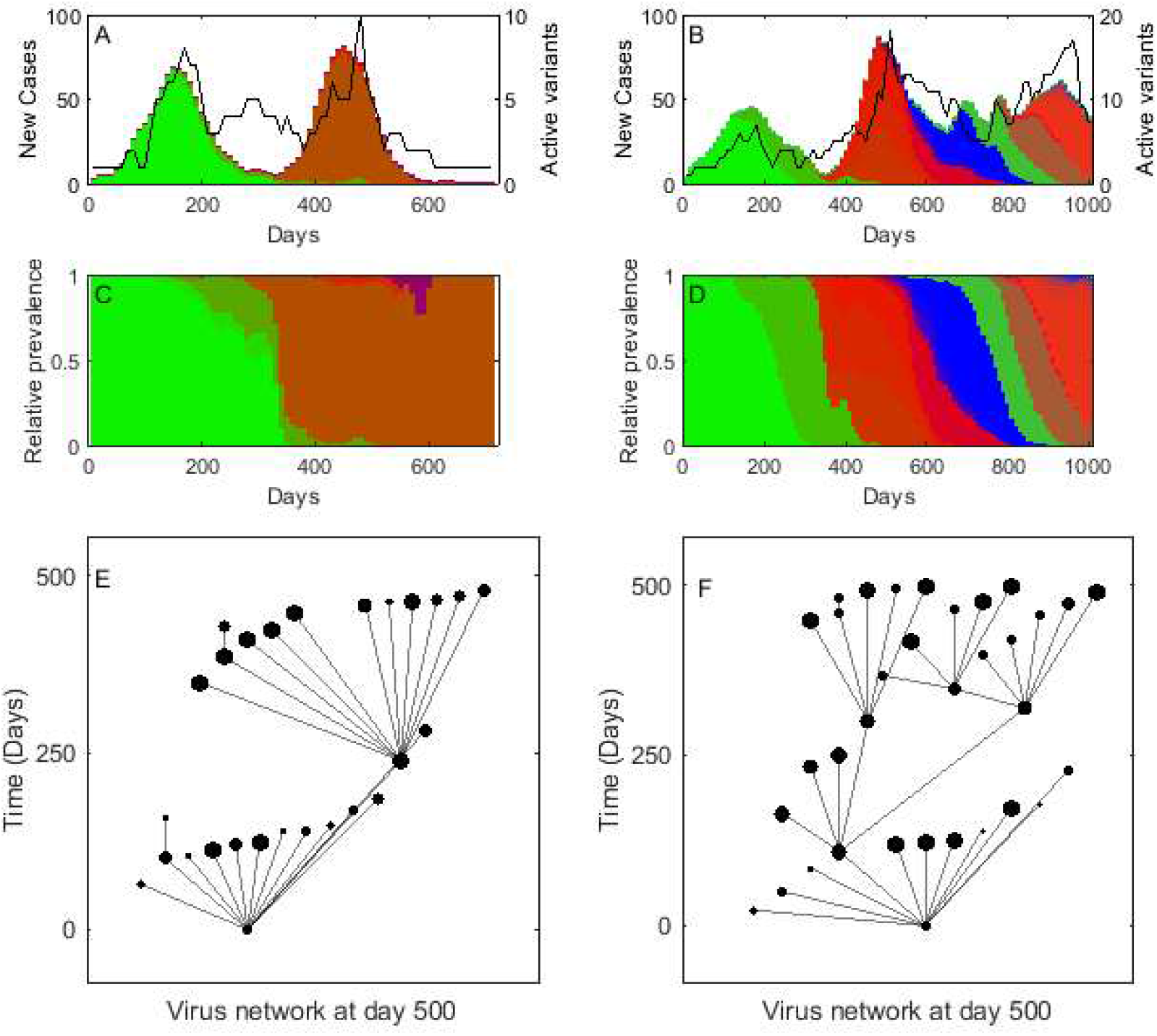
Model realisations either eliminate the virus or continue indefinitely with multiple variants. A and B show the number of new cases each day (averaged over a week, coloured bars) in two realisations of the model. Each virus variant is given a different bar colour. The black line shows the number of active variants (i.e. variants which had new infections) each week. C and D repeat the bars of A and B but scaled by the total number of new cases that week, to give the relative prevalence of each variant. E and F show the tree of variants at day 500, vertical position shows the time the variant appeared, node size shows the transmission rate *R*_*j*_ of each variant. The realisation in A, C and E eliminates after 650 days due to herd immunity. The realisation in B, D and F continues indefinitely. Bar colour is chosen randomly and is not related to any properties of the variant. Parameters as in Table 1 for high cross-immunity between strains and low transmission *p*_*mutate*_ = 0.005.

Initially, the second realisation (Figure 1 B, D, F) follows a similar pattern, the first wave peaking at day 150 is dominated by a single variant, the second wave peaking around day 450 is also dominated by a single variant. But as this second wave peaks more new variants become active and these variants are more distantly related than in the first realisation so there is less cross immunity between variants. Here after the second wave the disease becomes more widespread with a large number of active variants and elimination through natural herd immunity is no longer possible.

Extensive numerical exploration across many sets of parameter values shows that over 95% of realisations that eliminated have less than 200 variants in total. This was used to give a numerical definition of elimination, i.e. any realisation that reached 200 variants was presumed to continue indefinitely without elimination. In the scenarios below, simulations are run until either elimination is achieved or 200 variants have arisen whichever is sooner.

Figure 2 shows the probability of elimination with no vaccination for a range of mutation rates under the four transmission and cross immunity scenarios. In all scenarios as the mutation rate increases and new variants are more likely to appear the probability of elimination by herd immunity caused by infection decreases. This probability is higher if new variants are able to reach higher transmission levels, i.e. *R*_*max*_ is greater as there are fewer non-pharmaceutical interventions in place to keep transmission rates low. Similarly, if infection provides only a small probability of immunity to an evolved variant, i.e. there is low cross-immunity between variants, the probability of elimination is lower.

**Figure 2:**
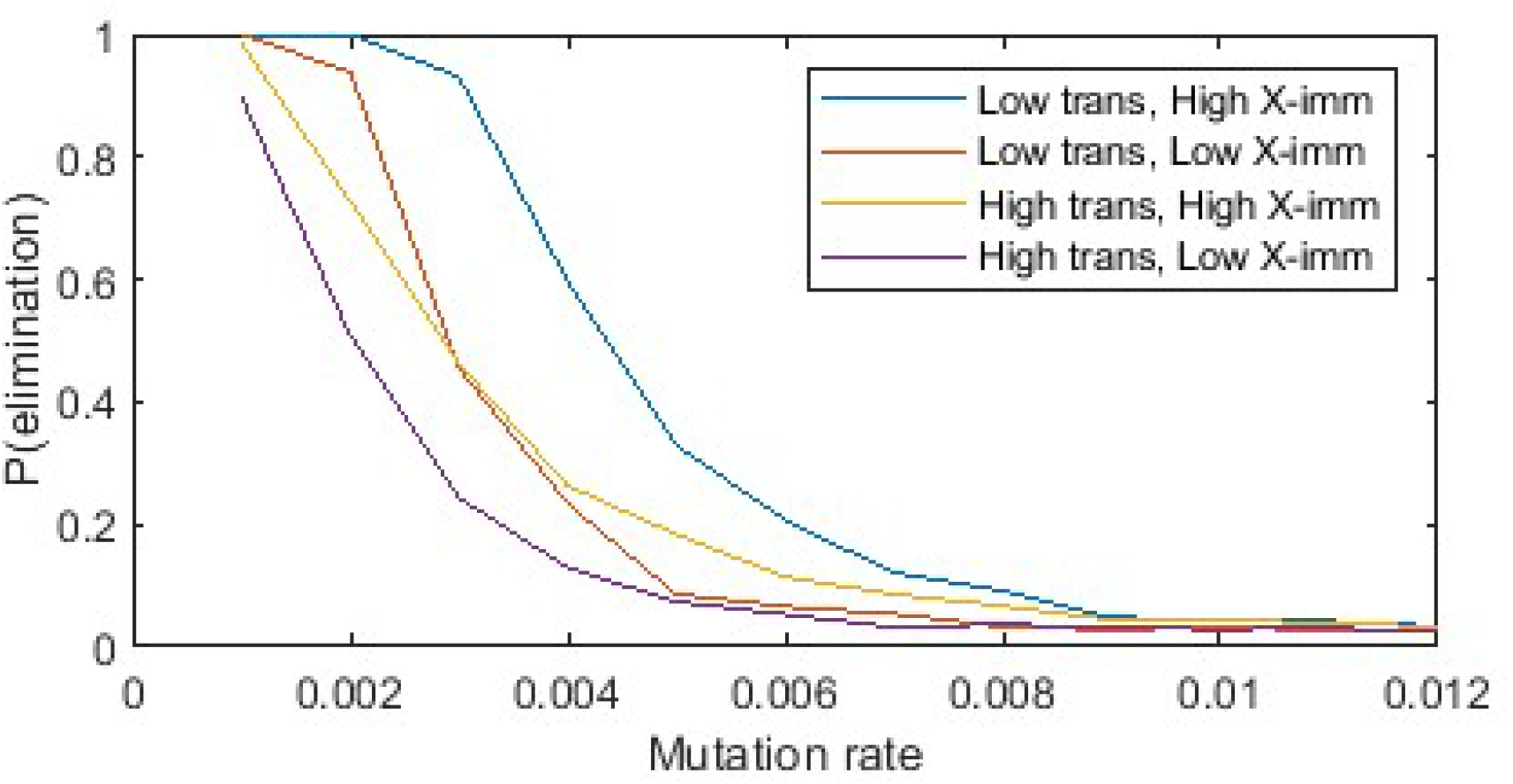
The probability of elimination decreases with mutation rate. The probability of elimination with no vaccine strategy. High transmission rates and low cross-immunity result in a lower probability of elimination. Calculated from 1000 realisations at each mutation rate, parameters as in Table 1.

## Results – Vaccines available

We now analyse the model results when a vaccine is available and can be updated to target new and emerging variants. We assume the first vaccine is available 350 days after the first infections and updated every *D*_*update*_ days after that. We consider a simple rolling vaccination programme where there is only one vaccine available at any time. Vaccine delivery is constrained by logistics, i.e. there is a maximum number of vaccine doses that can be administered each day. Every vaccine is 100% effective against the variant it was developed for and the immunity for other variants follows the immunity model. There is 100% vaccine take up, i.e. every individual in the population will take the vaccine if offered. We fix the mutation rate at 0.01 mutations per infection so with no vaccine the probability of elimination is almost zero. New vaccines are targeted at the variant that is most prevalent (i.e. has the most active cases) on the day the new vaccine begins delivery (i.e. there is no vaccine development lag), and that has not been a vaccine target before. Each day a fraction *f*_*V*_ of the population is vaccinated.

### Vaccine development speed

Figure 3 shows the results from each scenario for different vaccination rates, i.e. the fraction of the population that is vaccinated each day, and different updating rates. As the vaccination rate increases the probability of elimination also increases. Fast vaccination updates every 50 days are more effective in achieving elimination than slower updates of every 250 days. This improvement with update rate is most pronounced when the probability of achieving elimination is high. If the probability of achieving elimination is low, usually due to a slow vaccination rate, then more regular updating of the vaccine for new variants has only a marginal affect at best. High transmission rates have a very detrimental effect on the elimination probability. Given that the virus is eliminated, the proportion of people infected and the time taken to eliminate is not significantly affected by the vaccine update rate but is affected by a very slow vaccine delivery rate.

**Figure 3:**
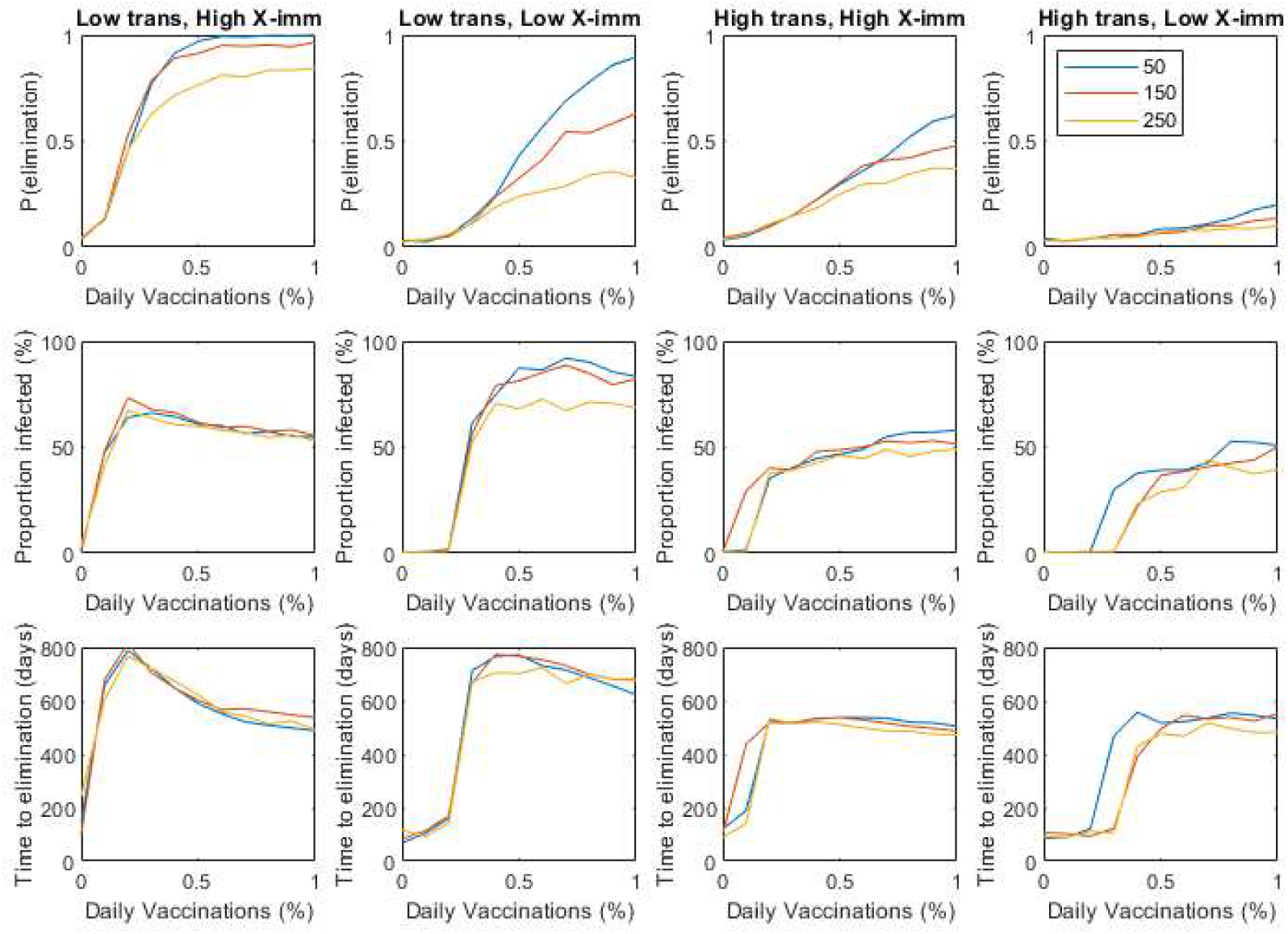
Faster delivery and faster updating of vaccines leads to a higher probability of elimination. The probability of elimination for a range of vaccination delivery rates, given as fraction of the population vaccinated each day, for three different vaccine updating rates (50, 150 and 250 days) across three different infection scenarios. Also the proportion of the population infected and time to elimination given elimination is achieved. Parameters are as in Table 1.

### Vaccine priority

We assume that there is a priority order for vaccine delivery. This order is decided on external factors such as job status (e.g. frontline workers and medical staff) or probability of severe disease. We assume these factors do not depend on an individual’s transmission rate, i.e. individual’s with higher transmission rates are not given higher priority, so do not affect the model presented here. If vaccinations are constrained by logistics, a simple question is: If a vaccine is updated before the entire population has been vaccinated who should receive the new vaccine? Should the programme continue and vaccinate previously unvaccinated individuals with the updated vaccine or should it reset back to the highest priority individuals, leaving some individuals completely unvaccinated but the higher priority individuals are more fully protected. Vaccine priority is the same for each vaccine.

In the Continuous strategy presented in Figure 3, the updated vaccine is given to the next individual on the priority list. This means lower priority individuals will likely receive the new vaccine first but every individual in the population will be vaccinated before the first individual receives their second vaccination, i.e. the higher priority individuals will only receive a vaccine update after all the other individuals have been vaccinated. We compare this to a Restart strategy where each new vaccine is given to individuals at the start of the priority list first, i.e. high priority individuals will receive every new vaccine as soon as it is available, while low priority individuals may never be vaccinated. Figure 4 shows that in all four transmission and cross-immunity scenarios the probability of elimination is slightly higher using a Continuous strategy and ensuring all individuals have been vaccinated rather than returning to the high priority individuals. This strategy may result in the highest probability of elimination but it would take a structured population model, not used here, to assess whether this strategy resulted in an increased risk to higher priority individuals who may have a higher mortality rate or likelihood of severe infection needing hospitalisation.

**Figure 4:**
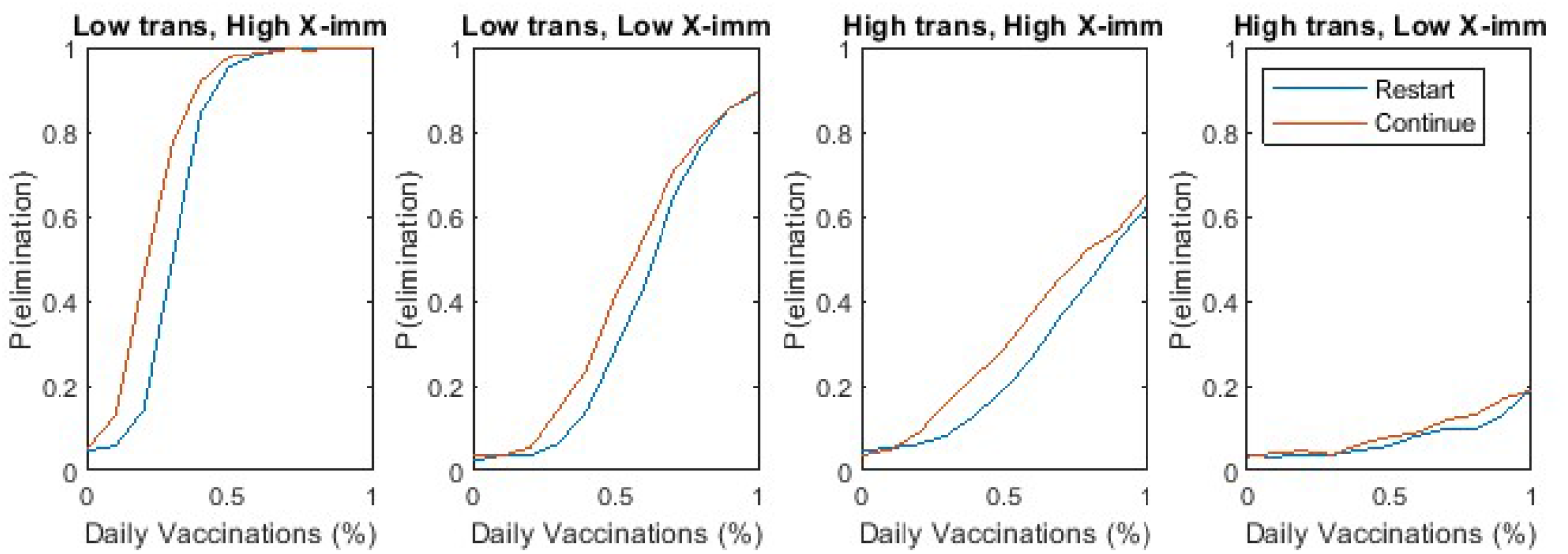
A continuous vaccination programme where updated vaccines are given to unvaccinated individuals, rather than revaccinating high priority individuals, gives a slightly higher probability of elimination in most scenarios. The probability of elimination for two different vaccination priority strategies as new vaccines are developed: Continuous – give the newly updated vaccination to the next person on the priority list to ensure everyone receives a vaccination; Restart – give the newly developed vaccine to the first person the priority list, i.e. some people get many vaccines and some get none at all.

### Vaccine uptake rate and effectiveness

Previous results assume that all individuals who are offered a vaccine will take it whereas Table S1 shows that in most countries uptake rates fall between 40% and 70%. As these rates are for the adult population and paediatric vaccines are often developed later than adult ones the actual uptake rate across the entire population may be lower still, especially in countries with a relatively young population. For example, in Israel almost 30% of the population are under 16 and ineligible for current vaccines (United Nations, 2019). In the low transmission scenario the maximum transmission rate of a variant is capped at *R*_*j*_ = 1.3 so using a very simple estimate of the herd immunity threshold (Murray, 2007) *H* = 1 - 1/*R*_0_ implies that provided 24% of the population was vaccinated there would be no epidemic. The corresponding threshold for the high transmission scenario (maximum *R*_*j*_ = 1.8) is 44%. Figure 5 shows the effect of uptake rate on the probability of elimination. If the uptake rate is significantly higher than the simple herd immunity threshold it has very little difference on the probability of elimination, however once the uptake becomes closer to the threshold the very high probability of elimination previously seen when the vaccine delivery is fast drops significantly.

**Figure 5:**
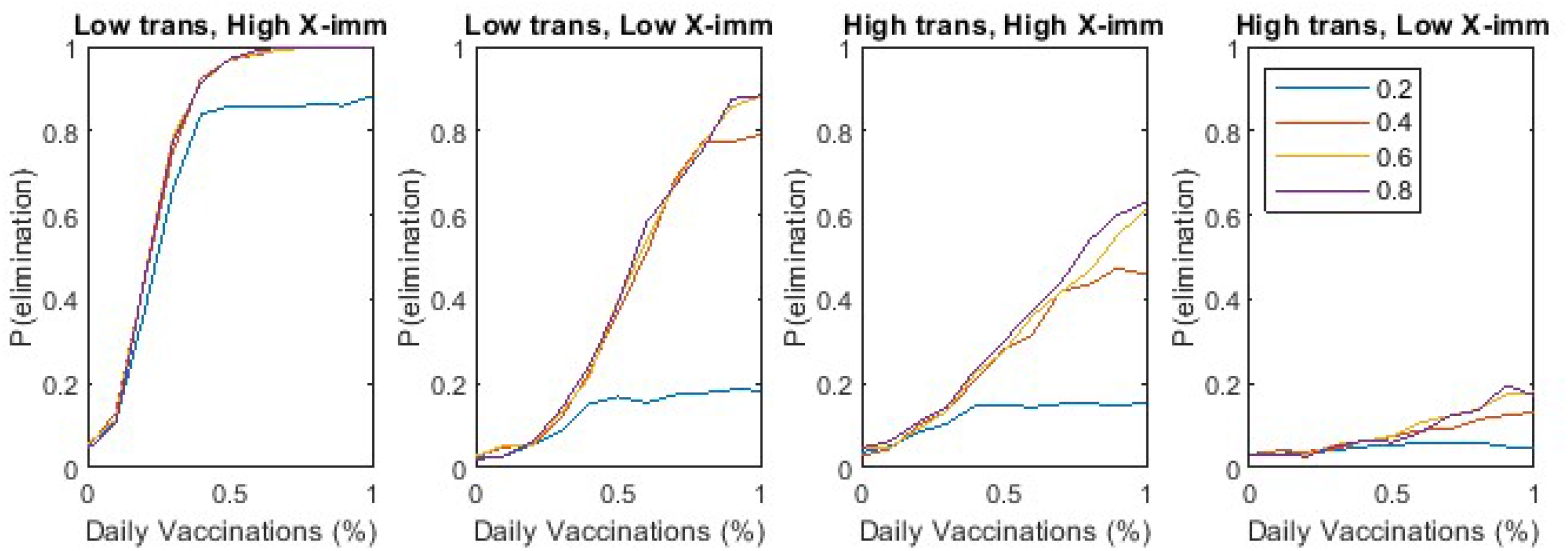
For higher vaccination rates if the uptake rate falls below the herd immunity threshold the probability of elimination drops. At lower vaccination rates, where the probability of elimination is always low it makes little difference. The probability of elimination at different daily vaccination rates for different vaccine uptake rates (20%, 40%, 60% and 80% of those who are offered the vaccine) in the four different scenarios.

### Population size

The results so far are shown for a relatively small population of only 10,000 individuals. Whilst this is computational cheaper it is less realistic. For the same reasons the results are constrained to relatively low maximum transmission rates. Figure 6 shows the output for a more realistic set of parameter values with population sizes up to 500,000, a higher maximum transmission rate *R*_*max*_ = 3, vaccine uptake rate of 60%, delivery rate of 1% of the population each day and vaccines are updated every 150 days. The threshold mutation rate at which elimination becomes considerably less likely decreases with population size but otherwise the qualitative results are very similar.

**Figure 6:**
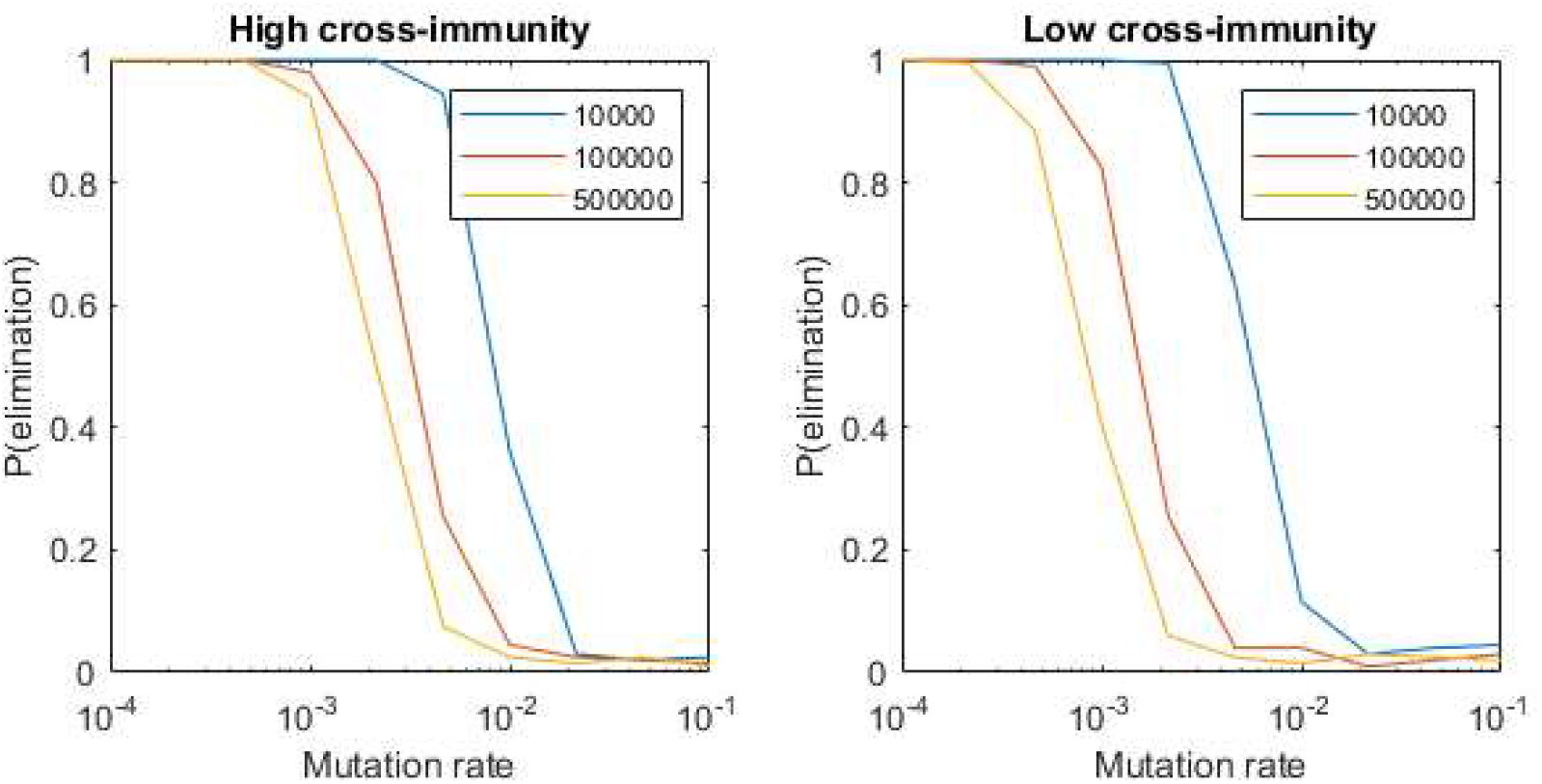
As the population size increases the model shows similar results with a slower mutation rate. The probability of elimination for different mutation rates for three different population sizes and two levels of cross-immunity.

### Predicting elimination with hard to estimate parameters

A key predictor of elimination is the rate at which significant new variants appear, *p*_*mutate*_. In practice predicting this rate and the level of cross immunity gained by infection or vaccination to particular strains is somewhere between tricky and impossible. Instead, using Monte-Carlo simulations across a wide range of parameter values (population size *N*_*p*_∼*U*(10^5^, 10^6^), mutation rate *p*_*mutate*_∼*U*(10^−4^, 10^−2^) and cross immunity *I*(1)∼*U*(0.1, 0.5)) we predict the probability of elimination, *P*(*elim*), conditional on the number of significant variants that have been observed after 300 days. We define a significant variant as one that has infected at least 100 individuals, i.e. it would likely have been spotted during standard levels of genetic sequencing. We use a general linear model of the form

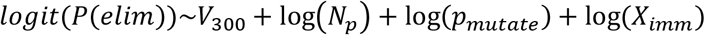

where *V*_300_ is the number of significant variants seen in that realisation during the first 300 days, *p*_*mutate*_ is the mutation rate, *N*_*p*_ is the population size and *X*_*imm*_ is the level of cross-immunity defined as a linear scaling on all the elements of *I*(*d*) in Table 2. Using all available predictor variables results in a highly accurate prediction of elimination probability (area under the receiver operator curve *AUC* = 0.97, Table 3). However, more encouragingly, using a simpler model with only the single predictor *V*_300_ gives a very good prediction of elimination probability (*AUC* = 0.94, Table 3).

**Table 3:**
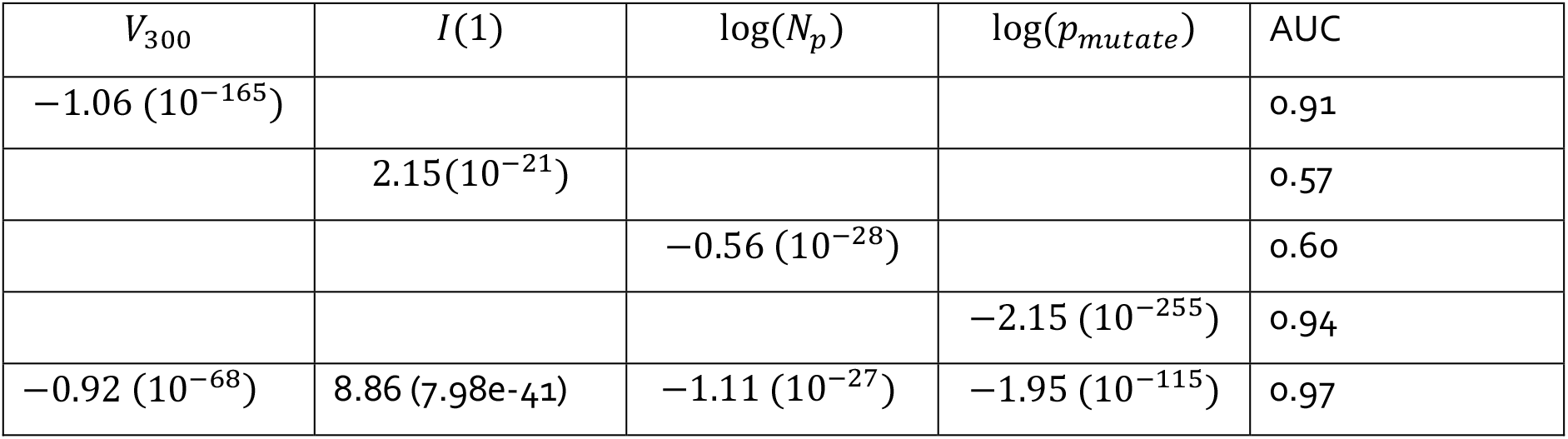
The observable variable *V*_300_, the number of variants seen after 300 days, is a very good single predictor of elimination probability. Coefficients (and p-values) of regression models predicting the number of significant variants and the probability of elimination.

The single variable model predicts that with no additional significant variants after 300 days, i.e. *V*_300_ = 1, there is an 86% probability of elimination. With 1 additional significant variant this falls to 68%. Using the more detailed predictor model shows that for any given number of additional significant variants the probability of elimination will decrease for bigger populations, lower cross-immunity and faster mutation rates.

Here we used the number of variants that arise in the first 300 days after the virus emerged. Very similar results can be seen for other time periods. When more than two or three significant new strains of the virus have emerged the probability of elimination drops significantly.

### Sensitivity analysis

We used a numerical definition of elimination where any realisation that reached 200 variants was assumed to continue indefinitely. This was checked extensively throughout the study and was seen to be correct in over 97% of realisations even for higher population sizes.

We have assumed a 100% vaccine effectiveness rate for the strain for which the vaccine was developed. Although we have not changed this explicitly the results of doing so would look very similar to the results for a lower uptake rate. For example the results for a 50% effective vaccine delivered at a rate of 1% of the population each day would be very similar to those for a 100% effective vaccine with an uptake rate of 50% delivered at a speed of 0.5% of the population each day.

We have assumed that the first vaccine to be developed takes approximately one year of development time. Explorations show that improving this initial development speed would increase the probability of elimination overall but the qualitative differences in results for changing other parameters would not change significantly.

Changing the disease transmission parameters, e.g. the number of infectious days does not change the overall results but does give a corresponding change in the timescale. Significant changes come with changing the transmission rates but these are explored through changes to *R*_*max*_, the maximum transmission rate.

Although cross immunity was varied in the results we assumed a linear scaling of cross-immunity across all distances in the virus tree. If cross-immunity remains high for closely related variants but decreases more rapidly with the network distance between variants this has a similar effect as lower overall cross-immunity.

Overall, the key results, in particular the usefulness of using the number of new variants after 300 days as a predictor of elimination, do not change if other model parameters are varied.

## Discussion

Our model shows that viruses which mutate into multiple new variants need fast vaccine delivery to be contained. Frequent vaccine updates to target new strains also increase the chance of containing the virus. Keeping transmission rates low through non-pharmaceutical interventions such as mask-wearing, social distancing and contact tracing will also improve the probability the disease is eradicated. Our model suggests that a continuous vaccination roll-out programme, where updated vaccines are given to unvaccinated individuals, rather than revaccinating high priority individuals, may slightly increase the probability of elimination. However, this prediction warrants further investigation using population-structured models to assess the risk this would pose to vulnerable individuals (e.g. frontline workers or those at higher risk of severe disease).

Under high vaccination rates, our results highlight the importance of ensuring that vaccine uptake rates are kept above the herd immunity threshold to avoid a considerable reduction in elimination probability. However, at lower vaccination rates this factor has less impact on the probability of elimination, which is low regardless.

Measuring the parameters needed to quantify models of multiple strains and predict the likelihood of long term elimination or even containment is somewhere between tricky and impossible. Instead we have presented a more observable quantity, the number of new variants that have caused a significant number of infections after a defined period of time, which is a good single predictor of the probability of elimination. As of May 2021 there had only been 5 countries that had seen a significant new variant emerge despite many countries having high infection rates. There has only been one nation that could claim to have had more than one new variant arise, i.e. India with very closely related strains of B.1.617. However, with each new strain that emerges the likelihood of long term containment decreases, especially as strains with very high transmission rates emerge.

However, when more than two or three new variants have spread to a significant number of people the probability of eliminating the disease drops significantly. At this point the long term outcome is similar to the current global influenza situation where the dominant viral strains change regularly, vaccines need to be constantly updated to stop the more severe outcomes and elimination is no longer an option.

## Data Availability

All data was previously published

https://data.worldbank.org

https://ourworldindata.org/covid-vaccinations

https://covid19dashboard.regeneron.com

https://github.com/YouGov-Data/covid-19-tracker

https://www.ipsos.com/sites/default/files/ct/news/documents/2021-03/global-attitudes-on-a-covid-19-vaccine-march-2021-

## Acknowledgements

This work was funded by the New Zealand Ministry of Business, Innovation and Employment and Te Pūnaha Matatini, Centre of Research Excellence in Complex Systems.

